# A preregistered, Open Pipeline for Early Cerebral Palsy Risk Assessment from Infant Videos

**DOI:** 10.1101/2024.11.06.24316844

**Authors:** Melanie Segado, Laura A. Prosser, Andrea F. Duncan, Michelle J. Johnson, Konrad P. Kording

## Abstract

Cerebral Palsy (CP), affecting approximately 1 in 500 children due to abnormal brain development, impacts movement control. Early risk assessment via the General Movements Assessment (GMA) at 3-4 months is highly predictive for CP but relies on trained clinicians. Machine-learning-based approaches for predicting GMA score from video have shown considerable promise, but typically rely on dataset-specific preprocessing, custom feature sets, and manually designed model pipelines, which make external benchmarking more difficult. This, combined with strict privacy constraints on sharing data, makes it challenging to train and evaluate models across datasets, which is important for assessing clinical utility. There is therefore a need to develop approaches that will work across different datasets to enable multi-site dataset aggregation and model training. To address this gap, we developed an end-to-end pipeline that uses off-the-shelf pose estimation, general-purpose feature extraction, and automated machine learning — none of which are tuned to a specific dataset. We applied this approach to a newly generated large dataset of 1053 infants (with approximately 10–12% positive class for adverse GMA outcome, drawn from a high-risk clinical cohort) within a preregistered study design. Model performance was evaluated on a strict “lock-box” test set, which remained untouched during any phase of model development or preprocessing optimization, and only used for evaluation once the final model and pipeline had been preregistered. The developed model achieved moderate predictive accuracy for clinician-assessed GMA scores (Area Under the Receiver Operating Characteristic Curve, ROC-AUC = 0.77; Area Under the Precision-Recall Curve, PR-AUC = 0.41). The moderate accuracy is noteworthy given the 10–12% positive class prevalence, and power-law scaling of ROC-AUC as a function of increasing dataset size. By releasing de-identified feature data and open-source code, and simplifying the training pipeline using AutoML, our work establishes essential groundwork for future robust, globally relevant CP screening tools suitable for low-resource settings.

**Key Points:**

- Introduced an open, accessible video-based pipeline, and used it to predict General Movements Assessment (GMA) scores (a key early indicator of Cerebral Palsy [CP] risk).
- Rigorously validated this pipeline on a large infant cohort (1053 videos), employing a preregistered design and a “lock-box” test set to ensure robust evaluation and minimize the risk of overly optimistic performance estimates.
- Demonstrated that relatively simple movement features, derived from handheld camera recordings achieve moderate predictive accuracy for GMA scores (ROC-AUC 0.77, PR-AUC 0.41), with power-law improvement with increasing dataset size, even under these stringent training and evaluation conditions.
- Designed the pipeline to facilitate broader application and collaborative research, particularly through its use of generalizable pose estimation and by enabling the extraction and sharing of de-identified movement features for aggregated dataset creation across clinical sites.
- Released the entire pipeline as open-source (including data processing, feature computation, and AutoML components) to promote transparency and reproducibility.

## Background

Cerebral Palsy (CP) is the most common cause of motor impairment leading to physical disability in children, affecting an estimated 2-3 out of 1000 infants globally [1]. In the USA alone, this results in approximately 1 million people living with impaired mobility due to CP at any given time, many of whom have lifelong disability. Early detection and rehabilitation before two years of age are critical, as beginning rehabilitation within this sensitive period for neural plasticity and motor development is associated with improved functional outcomes [2, 3]. Atypical movement patterns that indicate a high risk of developing CP are reliably detectable through a trained physician’s visual observation of movements at or before 10 weeks of age, but many infants are not evaluated by a physician until after severe overt motor impairments have developed. In practice, this means that CP is typically diagnosed between 6 and 24 months of age, which is near or beyond the end of the optimal window for intervention. There is therefore a need to develop automated early prescreening tools that can detect atypical patterns of motor development before they progress to more severe impairment, allowing for more efficient use of costly medical resources, and improved outcomes, particularly in low-resource settings.

CP risk is routinely assessed by clinicians based on visual observation of movements. One such assessment is the General Movements Assessment (GMA) [4], which is predictive of CP as early as 3 months of age based on expert classification of spontaneous infant movements. It distinguishes between *typical* and *atypical* General Movements (GMs), including the identification of Fidgety Movements (FMs) at 3-4 months, which are a precursor to coordinated, volitional movement. The absence of FMs at this age is 95% predictive of CP when combined with abnormal findings on brain MRI [2]. The GMA is typically scored from video and considers characteristics of movement quality, variability, and complexity. If these relevant movement features can be reliably computed from videos, it suggests that algorithmic approaches could feasibly be applied to estimate infant risk from movement features.

Numerous efforts by multiple research groups are underway to automate the GMA using computer vision and machine learning [5, 6, 7, 8, 9, 10, 11, 12, 13, 14, 15, 16, 17, 18]. These groups have all shown compelling evidence that GMA assessment, and by extension CP risk, can be predicted from video. However, how well existing approaches scale to new, unseen datasets remains an open question. Existing models often rely on hand-annotated or custom fine-tuned pipelines, and complete end-to-end implementations released for immediate external use are not widely available.

The advent of pre-trained vision transformers has enabled better feature extraction and multi-scale information fusion. This advance improves performance on data with joint- or limb-segment occlusions, as well as complex poses, both of which are common in spontaneous infant movement and challenging for infant pose estimation algorithms [19, 20, 21, 22, 23, 18, 24, 9, 25]. The combined advancements in computer vision, availability of human movement datasets, and development of open-source tools [26, 27] have significantly improved the reliability and accuracy of pose estimation and tracking outcomes [24, 19], even in challenging conditions. This raises the possibility that pre-trained vision transformers could be used for infant pose estimation without the need for custom finetuned models for specific datasets, overcoming a long-standing hurdle in the field[28, 24].

Most video-based automated risk assessment models are trained and evaluated on private datasets due to privacy constraints, which limits the ability of external groups to reproduce the full model behavior. When combined with the limited availability of complete model artifacts, it remains difficult to fully assess model generalizability across cohorts. These constraints also make it challenging to rule out optimistic performance estimates that can arise from methodological limitations or non-independent data splits [29]. For example, Gao et al. [6] trained a transformer model to detect fidgety movements using manually annotated video clips. This approach showed strong agreement with expert assessment, but the manual labeling required for training is resource-intensive, and the trained model artifact is not currently accessible. As a result, external groups would need to recreate these annotations and retrain the model in order to apply the approach to new cohorts, which makes it difficult for external groups to apply the method without recreating the training process. Similarly, Ihlen et al. [30] reported high sensitivity and specificity using backward prediction with a large set of handcrafted kinematic features. While this approach performed well, the reliance on a high-dimensional feature space makes it difficult to evaluate how the model behaves in independent cohorts [30, 31]. Groos et al. [9] demonstrated very strong performance on multi-site data using fine-tuned pose estimation and deep-learned representations, and although the method is well described, the full model artifacts are not currently publicly available, which limits opportunities for external replication.

Several prior studies have evaluated video-based CP risk prediction using external validation sets or fully held out test cohorts that were isolated from training and hyperparameter optimization [13, 9, 6]. These approaches follow established practice in machine learning evaluation for clinical applications. We adopt a preregistered lock-box approach that builds on these established practices. As in a conventional held out test set, the lock-box is used only for final evaluation. However, the lock-box adds an additional constraint, since it is evaluated only once within a prospectively defined analysis plan. The preregistered design differs from standard practice in that the evaluation dataset was specified in advance of any model development. This preregistration also fixes the preprocessing pipeline, model choices, and trained model prior to accessing the lock-box. For a single site study such as ours, this prospective specification increases transparency and reduces analytic flexibility [32]. To our knowledge, formal preregistration of a prospectively isolated test set remains relatively uncommon in this literature, and we provide our implementation (with code and data) as one possible template for future studies.

The models cited above and many others provide compelling evidence that GMA and overall CP risk can be predicted from video, but the diversity of methodological approaches and variability in what materials are publicly available currently make it challenging to assess clinical applicability across sites. Moreover, the lack of public datasets makes it difficult to compare across sites and train on multi-site datasets. Sharing videos and even keypoint time-series across clinical sites is prohibitive due to privacy and ethical constraints. In contrast, kinematic features and model weights can be shared publicly without any of these concerns and offer a simple, effective way to combine datasets and train models on more data. This is important given the low prevalence of atypical movement patterns in each dataset, especially when considering additional factors like CP subtypes and severity.

Our objective was to address these limitations by developing a pipeline, testing on a large dataset, and facilitating replication by other researchers. Accordingly, this study introduces and rigorously evaluates this open, generalizable pipeline, showcasing its performance on a large clinical dataset and its potential for enabling more accessible CP research. We assembled a large dataset of clinician-labeled videos from our institutions’ United States CP Early Detection and Intervention Network site’s data. To compute accurate movement features, we first selected an open-source pose estimation algorithm that performed well on our infant dataset based on clinical expert review. We then computed 38 features from the 2D pose estimates, describing posture, velocity, acceleration, left-right symmetry, and movement complexity. All 38 features were selected based on clinician-determined relevance to movement evaluation and preregistered prior to this study. To validate the pipeline, we trained a classification algorithm using automated machine learning to predict GMA scores.

We developed a pipeline for predicting infant CP risk from video using an off-the-shelf pose estimation algorithm, simple preregistered features, and automated machine learning that limits bias during hyperparameter optimization. We demonstrated that these movement features predict GMA scores in one of the largest infant datasets to date, and we released our feature dataset and the code needed for other researchers to process their own data, thus laying the groundwork for dataset sharing and collaborative model training (Figure 1).

**Figure 1.**
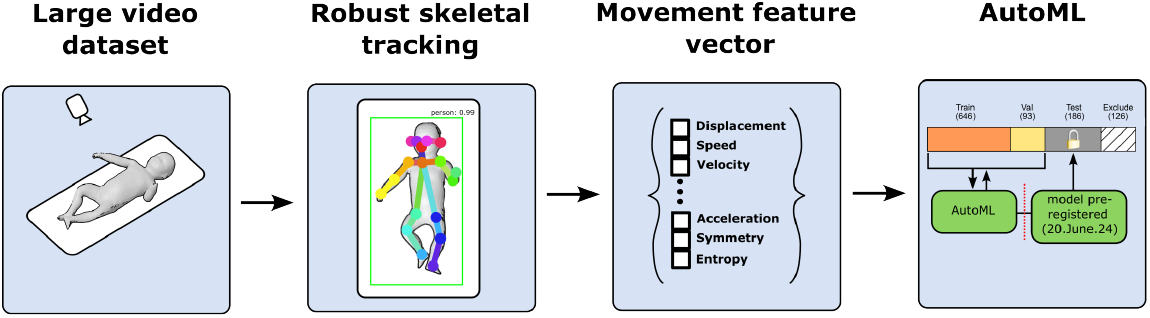
Process for rigorous evaluation of automated clinical score prediction. Each step of the model development process was preregistered, including subject IDs for each training split, pose-estimation algorithm selection, movement features, and AutoML model. Pose-estimation method was preregistered prior to feature computation. Features were preregistered prior to model training. Model was preregistered prior to testing on lock-box.

## Data description

### Collection of a large clinical dataset

Data were collected between May 2019 and December 2023 as part of standard clinical care by team members of the Children’s Hospital of Philadelphia (CHOP) site of the U.S. CP Early Detection and Intervention Network and entered into a REDCap database. This included the secure uploading of videos recorded on iPads or iPhones, GMA scores and demographic information. Access to this clinical database was restricted to hospital staff and authorized researchers. The GMA was administered in accordance with CHOP’s participation in the Cerebral Palsy Foundation’s Early Detection and Intervention network, which follows international diagnostic guidelines. For all infants who were between 10-20 weeks post-term age (corrected for preterm birth, if applicable) at the time of a clinic visit, and whose parents or legal guardians agreed to video recording for clinical care, clinicians captured a 1-2 minute video of the infant lying supine from a top-down perspective using handheld cameras. 10-20 weeks is the usual age for an infant’s first visit with the Neonatal Follow-up Program high-risk infant follow-up clinic, and a 1-to 2-minute video was deemed by two evaluators to be sufficient for GMA administration.

Infants were observed in minimal attire for unobstructed visibility of the trunk, shoulders, and extremities to facilitate the observation of natural movements (typically wearing a diaper only). The use of pacifiers, toys, or engagement in communication with the infant during the assessment was prohibited and other distractions that could potentially influence the outcome were minimized. If patients missed their clinic visit during this time period, parents were instructed on how to capture the video and provided a link to upload the video into REDCap.

### Video characteristics

The video dataset included 1053 recordings (one per infant), with a mean frame rate of 29.93 ± 3.28 frames per second (FPS). There were a few exceptions including videos at 15 FPS (3 videos) and 120 FPS (7 videos). All pose-data processing was normalized to each video’s frame rate.

The average number of frames per video was 3234 ± 762 for FM+ infants and 3447 ± 1031 for FM−. This corresponds to mean video durations of 112 ± 26 seconds for the FM+ videos and 119 ± 36 seconds for FM− videos. While the FM− videos had a slightly higher mean (7s) and greater variability in duration (10s), a Kolmogorov–Smirnov test indicated no significant difference between the distributions (p = 0.05), suggesting that video length is unlikely to bias downstream comparisons of movement patterns.

Orientation was highly uniform, with 1036 videos in landscape orientation and only 17 in portrait. Video resolutions were also largely consistent, indicating a standardized acquisition protocol suitable for motion quantification. 880 videos were collected at 1280×720 resolution, and a small number had other resolutions such as 480×272, 568×320, 1280×712, and 1920×1080. All video parameters are documented on the OSF preregistration site.

Camera angle can introduce geometric distortion that affects apparent bone lengths and, by extension, the accuracy of 2D joint angle calculations. To evaluate the reliability of joint-based metrics in this dataset, we estimated the average wingspan-to-body-length ratio as a proxy for viewing angle. Across the dataset, this ratio was 0.77 ± 0.12. The ratio was calculated by dividing a ‘wingspan’ proxy by a ‘body length’ proxy. The ‘wingspan’ was defined as the range of x-coordinates (max x − min x) and ‘body length’ as the range of y-coordinates (max y − min y), both derived from a comprehensive set of body joints (including arms, shoulders, hips, and legs – excluding head keypoints) after rotating each pose to a head-up orientation and normalizing torso length to one unit. While not a perfect metric, significant deviations from expected anatomical ratios would indicate non-top-down camera positions. The narrow variance in this measure supports the conclusion that the vast majority of recordings were obtained from a largely orthogonal top-down viewpoint, thereby minimizing projection errors and supporting valid kinematic analyses.

### Clinical evaluation

The evaluation process was characterized by the involvement of over 20 clinicians, including physical and occupational therapists, nurse practitioners, and physicians; several with additional advanced training. The GMA score (FMs present [FM+], absent [(FM−)] or abnormal) was determined after adjudication by two independent clinician reviewers. In instances where disparities in assessment arose, a third evaluator was consulted. Videos with uncertain scores were reviewed in weekly meetings convened by the site’s team.

This entire clinical scoring process was conducted entirely independently of the feature selection, data processing, and model development pipeline. This strict separation is essential as it helps ensure that our model’s performance was validated against objectively derived clinical labels, thereby minimizing potential for circular reasoning or bias that could artificially inflate its predictive accuracy.

### Patient characteristics

To assess how well we could predict GMA score from clinician-selected movement features in a large sample, we used videos that were collected as part of standard clinical care. In total there were 1053 videos from the Children’s Hospital of Philadelphia. The sample of 1053 infants was sex-balanced, with 55% girls, 45% boys, and 1% unknown/unspecified (Table 1 A). It also comprised a wide range of race/ethnicities, including White (38%), Black/African American (35%), ‘Other’ (10%). The remaining 16% of responses were spread across Multi-Racial, Asian, Indian, American Indian/Alaskan, Native Hawaiian, and Not Reported/Unknown/Other. Of reported ethnicities, 8% were Hispanic/Latino (Table 1 B).

**Table 1.** Demographic and race/ethnicity characteristics across Train, Val, and Test (lock-box) sets.

| (A) General Demographics |  |  |  |  |  | (B) Race and Ethnicity |  |  |  |
| --- | --- | --- | --- | --- | --- | --- | --- | --- | --- |
| Feature | Train | Val | Test<br>(lock-box) | $p_{T-V}$ | $p_{T-T}$ | Category | Train | Val | Test (lock-box) |
| Sample Size (N) | 646 | 93 | 186 | — | — | % White | 38.5 | 39.6 | 39.0 |
| % Female | 54.6 | 55.9 | 55.1 | 0.9 | 0.9 | % Black or African American | 33.6 | 32.3 | 32.6 |
| Gestational Age (days) | 220.4 | 224.4 | 224.6 | 0.2 | 1.0 | % Other or Multiracial | 18.7 | 18.3 | 19.3 |
| Chronological Age (weeks) | 24.6 | 21.9 | 22.2 | 0.2 | 0.7 | % Not Reported (Race) | 9.2 | 9.8 | 9.1 |
| % FM- | 10.8 | 10.8 | 10.2 | 0.9 | 1.0 | % Hispanic/Latino | 8.1 | 7.5 | 7.5 |
|  |  |  |  |  |  | % Not Reported (Ethnicity) | 7.9 | 8.6 | 9.1 |

Demographic and race/ethnicity characteristics across Train, Val, and Test (lock-box) sets. $p_{T-V}$ and $p_{T-T}$ indicate comparisons between Train vs. Validation and Train vs. Test (lock-box) sets, respectively. In panel (A), $p$ -values were obtained using $\chi^2$ tests for categorical variables and two-sample $t$ -tests for continuous variables. Race and ethnicity reflect self-reported categories. Percentages may not total 100% due to rounding or non-exclusive identification.

This cohort includes a high proportion of infants with known risk factors for neurodevelopmental delay. Specifically, 70% (n=653) were born preterm (<259 days gestation), 45% (n=419) had very low birth weight (<1500g), and 28% (n=265) met criteria for extremely low birth weight (<1000g) [33].

Each of the video recordings used for analysis was determined evaluable by the clinical reviewers. In cases where infants were distracted during the recording session, a second video was obtained. Only the videos used for GMA scoring were included. The mean corrected age was 14.6 weeks (*±* 2.1 weeks). Of the 931 infants that remained after applying exclusion criteria (see: Inclusion and exclusion criteria), 826 were scored as having FM+ and 99 infants were scored as FM−. The remaining six infants were scored as having atypical fidgety movements (atypical FM) and were excluded from model training, but still included for pose-estimation and feature computation. The full list of excluded IDs can be found on the OSF preregistration site (https://osf.io/gztmd/) [34].

### Inclusion and exclusion criteria

For children still hospitalized at the time of the fidgety-aged GMA, Early Detection Team members captured the videos in the hospital as part of standard care. Exclusions were applied to intubated patients, those under the influence of sedation medications, within a week post-operative, on ECMO support, or diagnosed with myelomeningocele. The full dataset comprised 1053 infants. 122 were then excluded for meeting one or more exclusion criteria listed above.

Six infants with atypical FM were excluded from the Train, Val, and Test (lock-box) datasets, since there were not enough infants in this group for multi-class training and prediction (Figure 2). The remaining 925 videos were split into an analysis set (Train: 646, Val: 93) and a lock-box set (Test: 186), each containing approximately 10–12% of the FM– movement type. Note that the AutoML framework implements successive halving with internal cross-validation within the training data for hyperparameter optimization and model selection; the model-development Val set was used only for evaluation of model performance and was not involved in tuning or selecting models, while the preregistered Test (lock-box) set was reserved exclusively for the final evaluation.

**Figure 2.**
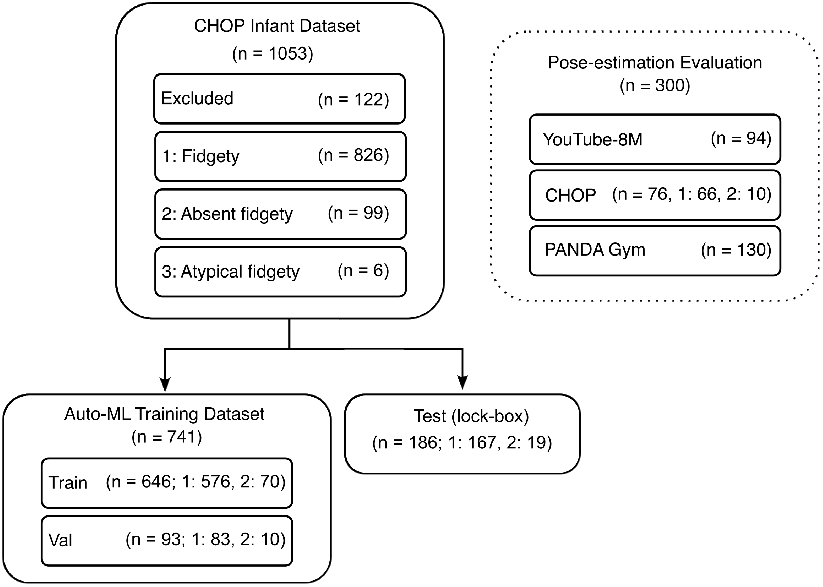
Datasets used for pose-estimation algorithm selection, AutoML training and testing, and lock-box evaluation. The pose-estimation evaluation dataset included 94 videos from the YouTube-8M dataset, 76 videos from the training subset of the CHOP Infant Dataset, and 130 additional videos of infants from ongoing projects in the lab. The CHOP Infant Dataset included 1053 infants; 122 were excluded for meeting one or more medical exclusion criteria prior to any analysis. Data were split into Train (646), Val (93), and preregistered Test (lock-box; 186) subsets, each containing approximately 10–12% of the positive class (FM–). The lock-box subset was isolated prior to pose-tracking algorithm selection and movement feature computation and was evaluated exactly once following preregistration of the final model and analysis plan. The six atypical-fidgety videos were excluded from model training but are included in the released feature dataset.

The splits were stratified to preserve the ratios of male and female infants, as well as age, and race and ethnicity, and chronological age on upload date (Table 1 A). There was a total recording duration of 60-120 s per infant.

## Analyses

### Developing a pipeline for robust skeletal tracking

#### Performance and validation of skeletal tracking pipeline

Infant videos pose unique challenges for pose-estimation algorithms due to frequent irregular body poses, the presence of body-like objects (e.g., toys or cartoons), high levels of self-occlusion, and different body proportions relative to adults [28]. Historically, algorithms such as OpenPose [35] fail in such conditions, leading to unreliable pose estimates [22, 9, 24, 18]. Extensive fine-tuning is often required to improve accuracy on each individual infant dataset [36, 24], making it difficult for researchers without the time and technical skills to benefit from custom models. Typically, each research group will annotate a subset of their own data to fine-tune a model that works well for their specific dataset. However, at the time of algorithm selection, the fine-tuned models referenced in the literature were not yet available in fully packaged form suitable for immediate external testing. Increasingly this is no longer the case as open fine-tuned models for infant pose-estimation continue to be released and updated [18, 24].

One of the challenges in fast-moving fields like computer vision is the rate at which new models are developed. The open-source MMPose framework we used for the pose-estimation step of our pipeline simplifies testing multiple algorithms with different weights and adopting new ones as they are released [26]. We first tested various pre-trained algorithms on a diverse infant-video set (Figure 2). We then compared their performance to our lab’s previously fine-tuned OpenPose model [5]. As we did not have a ground-truth annotated subset of videos to benchmark against, we relied on feedback from experts trained in scoring the GMA regarding whether or not the skeletal tracking was sufficiently good that a trained human would be able to administer the GMA on animated videos of the keypoint data. We found that the pretrained ViTPose-H [19] performed better than other pretrained models (HRNet, PVTv2) [37, 38] and our previously fine-tuned OpenPose model [39, 35, 40, 5]. Because our goal was to identify pose estimators likely to generalize across datasets without retraining, we evaluated all models in their pretrained form. ViTPose-H nonetheless achieved the highest tracking quality (as evaluated by our clinical experts) without any dataset-specific fine-tuning (Figure 3).

**Figure 3.**
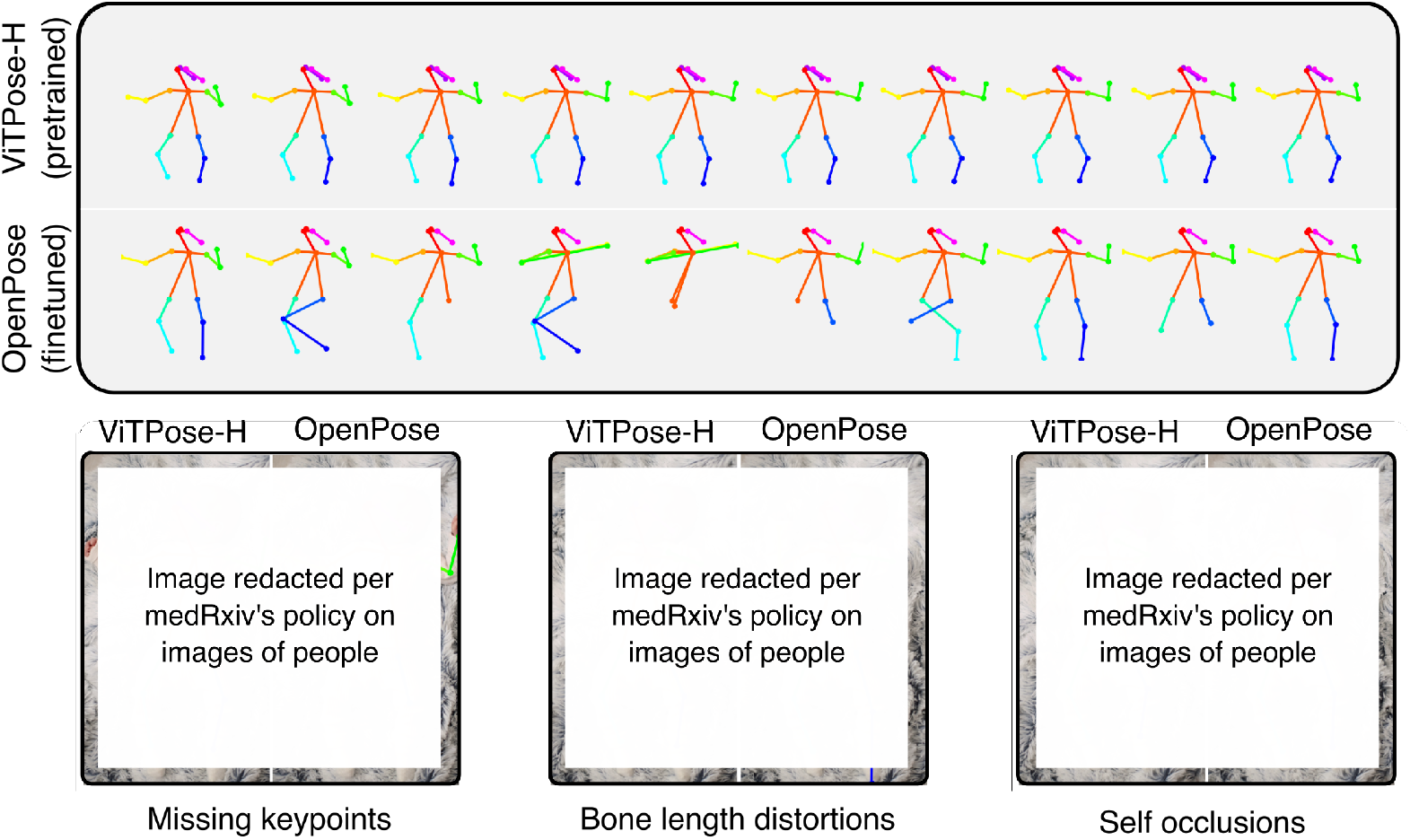
Improvements in skeletal tracking with pre-trained vision transformers. (Top panel) ViTPose-H (top row) produces consistent keypoint detections across frames in contrast to older algorithms like OpenPose (bottom row). (Bottom panel) Transformer-based approaches, such as ViTPose, learn adult skeletal priors and can infer missing keypoints (left), estimate bone lengths (center), and resolve self-occlusions (right) in infants.

We have made it as easy as possible for researchers to perform pose estimation on their videos by using a method and algorithm that we provide as a Docker container, built on open-source code that also provides its own Docker container. The original ViTPose-H weights are publicly available on HuggingFace from the ViTPose-H authors, and another group has released ViTPose models finetuned for infant pose estimation, with weights published on Zenodo [24]. In our pose-estimation code, users can specify any of these pretrained weight files through a simple configuration option.

#### Generalizability testing

The robustness and generalizability of ViTPose-H were validated through iterative review of pose estimates by clinicians trained in the GMA. We relied on expert judgment to determine whether the skeletal tracking was sufficiently robust for a clinician to assess fidgety movements from the keypoints alone.

To evaluate performance on videos beyond our primary dataset, ViTPose-H was also tested on two fully out-of-sample infant datasets from a separate project in the Rehabilitation Robotics Lab (“PANDA Gym”, n = 130; ages = 0-6 months, typically and atypically developing, 6 camera angles)[41], as well as on a set of 94 infant videos from the YouTube 8M dataset [5]. The algorithm produced consistent results across datasets (i.e., results that clinicians deemed to be sufficiently smooth), supporting its generalizability. To facilitate further testing and development, we have publicly released the ViTPose-H keypoints for the YouTube-8M videos. We propose that ViTPose-H offers a scalable and reliable solution for converting infant videos into skeletal tracking data without the need for fine-tuning, enabling broader applications in infant movement analysis.

#### Benchmarking against deep-learning models

To establish a baseline for how simple kinematic features perform relative to deep learning methods, we first surveyed openly available code from recent papers on automated GMA. For many recent papers, publicly released repositories did not contain all components needed for full replication, such as pretrained weights or complete preprocessing pipelines. Direct requests for model artifacts and evaluation data did not lead to access at the time of our study, which limited our ability to perform external testing.

One available implementation (STAM: Spatio-temporal Attention-based Model) [13] was complete and testable, and it initially appeared to show excellent performance (ROC-AUC = 0.86). However, we observed that the available implementation used record-wise splitting. This practice is a well-documented source of overfitting in medical data [29]. After adjusting the split to avoid overlap, performance was substantially lower with an ROC-AUC of 0.60. By contrast, our pipeline achieves a substantially stronger ROC-AUC of 0.77 on the identical preregistered data splits.

This finding underscores a broader point that progress in the field depends on open data and code sharing together with rigorous evaluation practices that minimize overfitting.

### Feature relevance and clinical interpretation

A set of 38 kinematic features was selected based on clinician input [34, 42], designed to capture the displacement, velocity, acceleration, and entropy of key body parts: wrists, ankles, elbows, and knees (Table 2).

**Table 2.** Summary of position and angular features.

| Position Features X & Y (ankles, wrists) |  |
| --- | --- |
| Cross-Correlation | Bilateral coordination/symmetry. |
| Entropy | Movement variability and complexity. |
| IQR Acceleration | Captures abruptness of movement. |
| IQR Position | Indicates range of movement in position. |
| IQR Velocity | Measure of movement smoothness. |
| Median Position | Captures postural biases or asymmetries. |
| Median Velocity | Typical speed of movement, reflecting consistency and effort. |
| Angular Features (knees, elbows) |  |
| Cross-Correlation | Bilateral coordination/symmetry. |
| Entropy | Movement variability and complexity. |
| IQR Angular Acceleration | Proxy measure of spasticity/stiffness. |
| IQR Angular Velocity | Measure of movement smoothness. |
| Mean Joint Angle | Deviations may indicate abnormal tone. |
| Median Angular Velocity | Measure of rigidity. |
| Stdev Joint Angle | Reduced variance may signal restricted movement. |

GMA-trained clinicians chose these 38 movement features because they are essential components of visual GMA scoring [4], though it is important to note that these are general kinematic descriptors that apply across contexts and age groups. We excluded features directly related to GMA-specific FMs to capture general movement patterns, enabling future work on earlier risk prediction, CP subtype classification, and other movement disorders.

### Model performance and validation

#### Generalizability and robustness of feature vector for risk prediction

Our initial analysis found considerable overlap in the 38 selected features for infants with and without fidgety movements (Figure 4),

**Figure 4.**
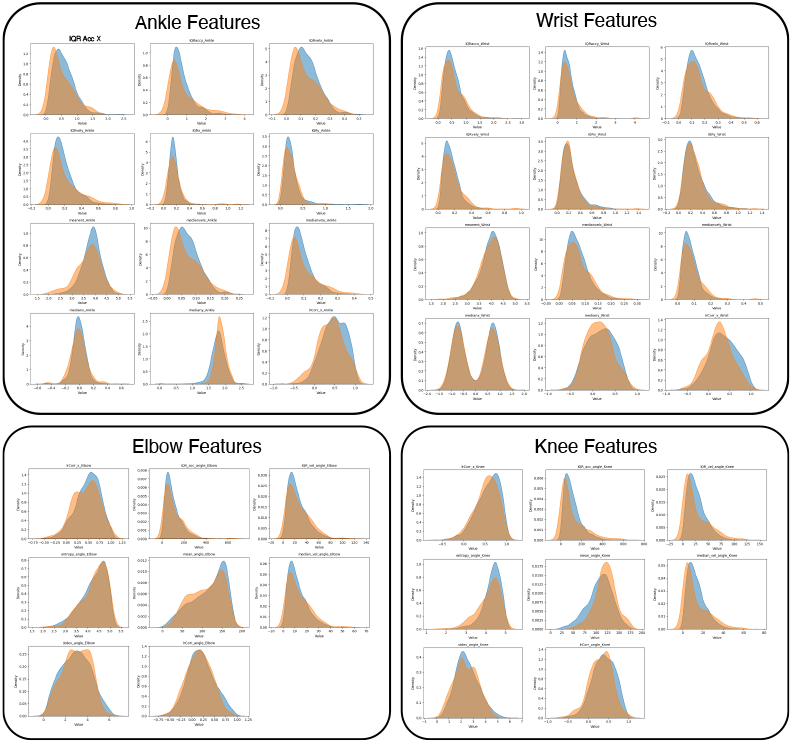
Individual feature distributions are highly correlated. Clinician-selected features, including XY features of the wrists/ankles and angular features of the elbows/knees, which are typically used for human assessment of risk are highly overlapping for FM+ (blue) and FM− (orange) movements, with no individual feature clearly predicting GMA score.

Table 2. Summary of position and angular features suggesting that no individual feature could differentiate high-risk infants. However, the aggregated feature vector was sufficient to predict GMA scores. Using the feature vector, the model achieved an ROC-AUC of 0.72 on the Val set (i.e., the preregistered Validation set of 93 IDs to be used for AutoML model development), and 0.77 on the Test (lock-box) set (Figure 5). Five-fold cross-validation, repeated with six random seeds (shuffling IDs from the Train/Val), yielded an average ROC-AUC of 0.73 ± 0.05. No IDs from the Test (lock-box) set were included in this cross-validation analysis. This demonstrates the model’s robustness and internal generalizability.

**Figure 5.**
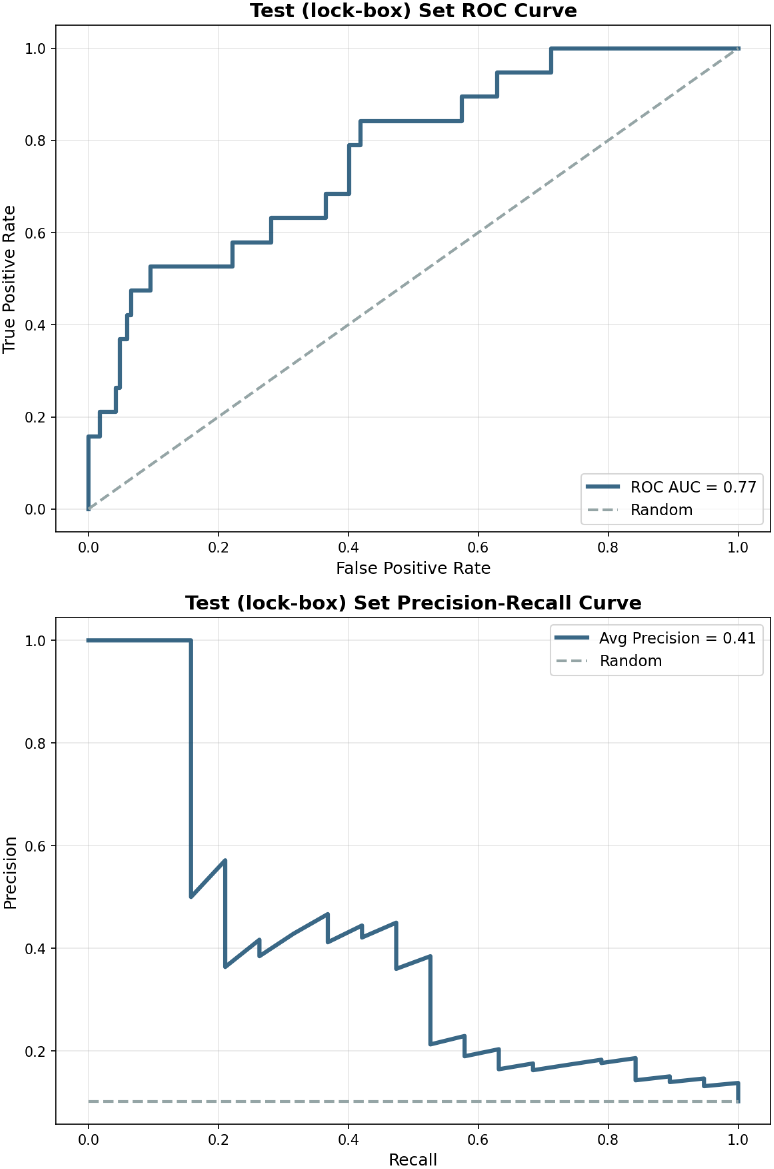
Model generalizes to lock-box set. Classifier trained on clinician-selected features using vanilla Auto-sklearn 2.0 shows a high ROC-AUC of 0.77 (Top) and Precision-Recall of 0.41 (Bottom) on lock-box set of 186 infants, having 10% representation of FM−. True positive rate is equal to the Sensitivity of the classifier, False positive rate is equal to 1-Specificity.

We also computed the Precision-Recall Area Under the Curve (PR-AUC), a metric particularly informative for imbalanced datasets typical of clinical screening contexts. Given the positive class prevalence of approximately 10–12% in this lock-box sample (FM−), the PR-AUC of 0.41 substantially exceeds the random chance baseline and highlights the model’s utility in identifying at-risk infants.

#### Illustrating sensitivity–specificity trade-offs in classification

The model produced continuous probability scores reflecting the likelihood of FM–. However, classification requires selecting a threshold. Threshold selection was not part of our preregistered model development plan, but to illustrate the tradeoff between sensitivity/specificity at a specific decision threshold, we selected the point on the Test (lock-box) ROC curve that maximized the difference between the True Positive Rate (TPR) and False Positive Rate (FPR). This threshold was defined as:

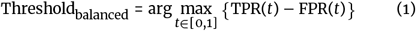

This corresponds to the threshold that maximizes Youden’s J statistic (*J =* sensitivity *+* specificity *– 1*), ensuring balanced sensitivity and specificity in the context of class imbalance.

The point that maximizes Youden’s J statistic on the Test (lock-box) ROC curve (Threshold = 0.61) yields a TPR of 53% (10/19) and a FPR of 10% (16/167). Samples with a model score above this value were classified as positive (FM−), and those below were classified as negative (FM+) (Figure 6). For comparison, the corresponding point on the Val set (Threshold = 0.26), yields a TPR of 55.0% (11 / 19) and a FPR of 24.6% (41 / 167) on the Test (lock-box) data, highlighting the importance of evaluating threshold stability across independent datasets to assess the robustness of model calibration and generalizability.

**Figure 6.**
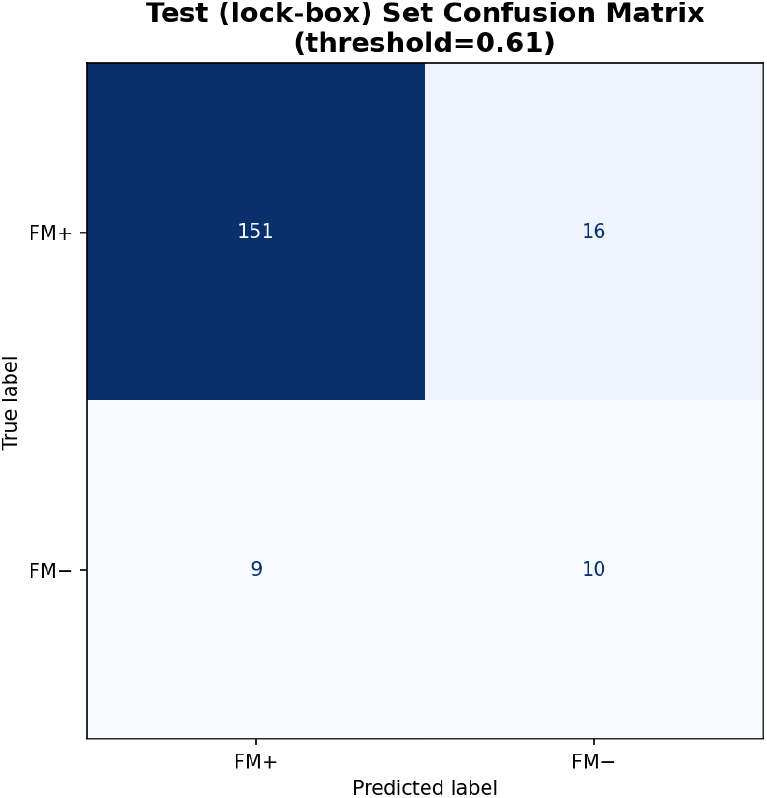
Confusion matrix for model predictions. Confusion matrix on the preregistered Test (lock-box) set, visualized at the operating point corresponding to the “optimal” threshold given by maximizing Youden’s J. This point was selected on the ROC curve for illustrative purposes only, to demonstrate the trade-off between sensitivity and specificity.

Importantly, the confusion matrix (Figure 6) is presented for illustrative purposes only. Our evaluation focuses on threshold-independent model performance, as quantified by the area under the ROC-AUC, rather than on classification accuracy at any specific operating point. Selecting a classification threshold after inspecting the ROC curve carries a risk of optimistic bias, and in future work we will include threshold selection in prospective analysis plans and preregister it alongside the model prior to evaluating on the test set.

#### Model training with Auto-sklearn 2.0

Experimenter-driven hyperparameter optimization and model selection are significant sources of bias in machine learning, often leading to models that don’t generalize well [32]. Automated Machine Learning (AutoML) frameworks like Auto-sklearn 2.0 address this by abstracting these decisions [43, 44]. Instead of manual tuning, Auto-sklearn 2.0 systematically explores algorithm choices and hyperparameter settings, leveraging meta-learning—insights from previous experiments (i.e., other, fully independent datasets with similar statistical properties) to interpret the characteristics of the input data, such as class imbalance. This allows it to strategically select appropriate handling techniques like SMOTE, class weighting, and appropriate evaluation metrics (e.g., stratified crossvalidation).

Auto-sklearn 2.0 uses Bayesian optimization to search model configuration spaces and can automatically construct ensembles of top-performing models. However, in the strict “vanilla” Auto-sklearn 2.0 configuration [44], both the training time (1 hour) and ensemble size (one model) are constrained. Although ensembles can improve robustness through variance reduction, they can also overfit when model selection repeatedly adapts to validation results as is common behavior in AutoML [43, 44]. We therefore used single-model training to obtain conservative, reproducible performance estimates and to preserve the integrity of the preregistered lock-box evaluation.

#### Rigorous methods to prevent overfitting

To ensure that the model’s performance was as unbiased and generalizable as we could achieve using data from only one site, a lock-box test set of 186 infants (19 FM−) was randomly selected before model training and preprocessing optimization. This lock-box dataset was only accessed after preregistering all features, preprocessing steps, and algorithms. Evaluating on the lock-box set yielded a ROC-AUC of 0.77, and the precision–recall curve showed a PR-AUC of 0.41 (Figure 5). The ROC-AUC was closely aligned with the crossvalidation performance indicating minimal overfitting. While both measures are lower than clinician performance reported in the literature and that of many other ML models, they are notable given the simplicity of the features and the stringent training parameters.

This rigorous validation suggests that our pipeline generalizes well to unseen data collected under similar conditions. It provides a reliable, reproducible approach for training models to identify infants at high risk for CP.

#### Scaling analysis

To demonstrate the effect of aggregating simple features, we performed a scaling analysis. We took the complete feature dataset and split it into training sets containing 50, 100, 200, 400, and 800 infants, using the same test set of 100 infants (12% FM–). We found that the improvement in ROC followed a power-law relationship, as indicated by the approximately linear trend in the log-log plot (Figure 7).

**Figure 7.**
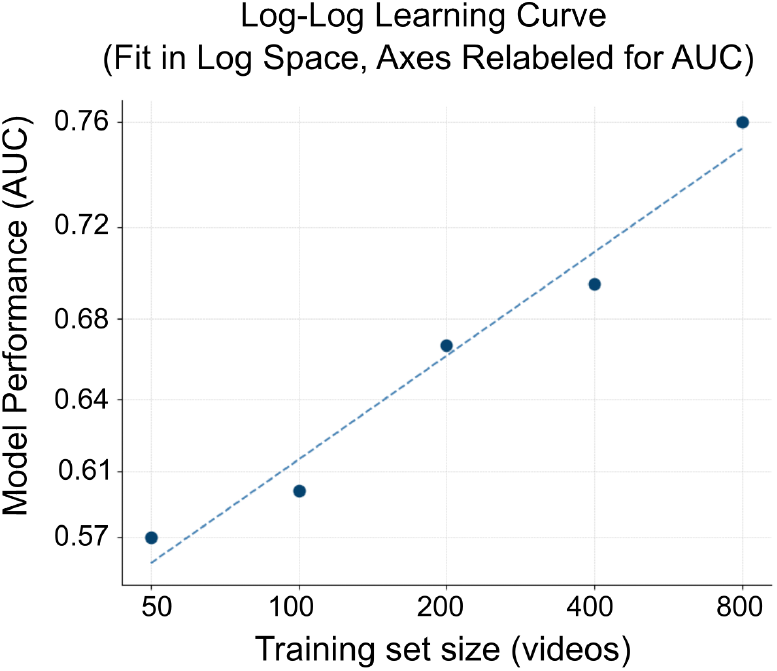
Improvements in model performance as a function of training-set size. The relationship between training size and performance was modeled in log–log space log(*n*_*videos*) vs. log(*1*−*AUC*), which follows a power-law trend. For interpretability, the x and y axes have been re-labeled to show the corresponding linear AUC values rather than log(*n*_*videos*), log(*1*−*AUC*). All AUC values represent the average of 6-folds cross-validated training results.

#### Explainability and feature importance

To provide some measure of model explanation, we implemented a standard permutation importance test using the sklearn framework [44, 45]. This approach systematically permutes feature values to assess the extent to which model performance depends on each input. We found that knee and elbow features contributed most strongly to model performance, followed by the wrists and ankles (Table 3). However, features with measurable importance were distributed across all body parts and statistical descriptors (mean, IQR, standard deviation) (Figure 8). This is consistent with established understanding of GMA assessment, which relies on global, whole-body movement patterns rather than isolated features.

**Table 3.**
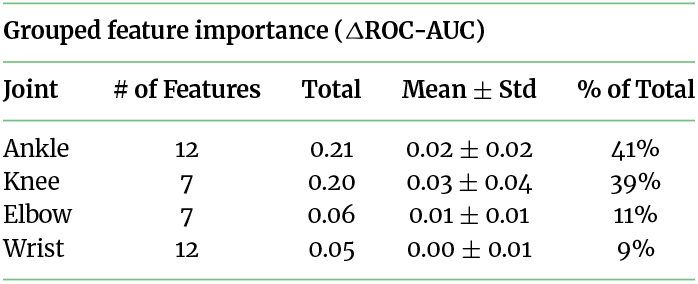
Grouped feature importance by joint.

| Grouped feature importance ($\Delta\text{ROC-AUC}$) | | | | |
| --- | --- | --- | --- | --- |
| Joint | # of Features | Total | Mean ± Std | % of Total |
| Ankle | 12 | 0.21 | $0.02 \pm 0.02$ | 41% |
| Knee | 7 | 0.20 | $0.03 \pm 0.04$ | 39% |
| Elbow | 7 | 0.06 | $0.01 \pm 0.01$ | 11% |
| Wrist | 12 | 0.05 | $0.00 \pm 0.01$ | 9% |

**Figure 8.**
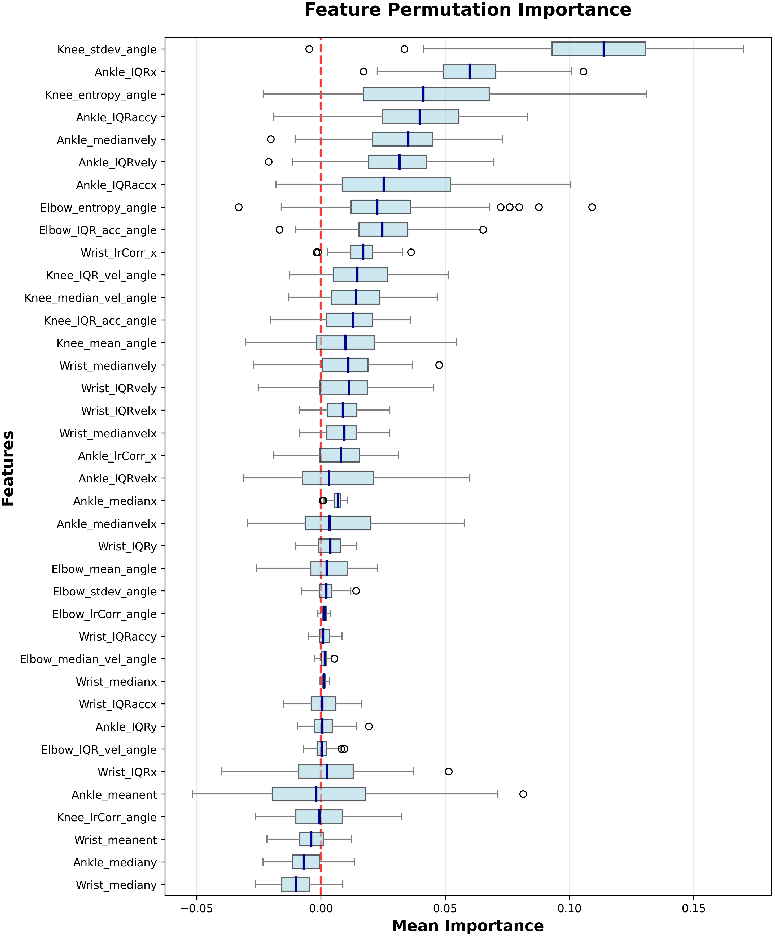
Permutation importance for each feature. Individual feature importance is low and highly variable, indicating that the model is using a combination of all features for predictions.

At the same time, we emphasize that this type of analysis has important limitations. Because many features are highly correlated, their contributions cannot be cleanly disentangled. Moreover, placing too much weight on individual features, or using them as the basis for feature selection, risks overfitting and may reduce generalizability. As our scaling experiments demonstrate, model performance improves most reliably when aggregating across increasing sample size, rather than prioritizing any one feature set. Thus, while this analysis provides some intuition about which features may be more influential, we caution against overinterpretation and underscore that robust performance derives primarily from data breadth rather than feature selection.

## Discussion

Here, we present an open, preregistered pipeline that was developed for video-based movement analysis and subsequently evaluated on the task of predicting GMA scores (a strong indicator of CP risk) using rigorous methods. We used an exceptionally large sample (training set: 646; overall >1,000 infants), a simple, generic movement-based feature vector, and preregistered each step before testing on a Test (lock-box) set of 186 infant videos. We found that our algorithm, trained using this pipeline, performs well (ROC-AUC = 0.77, PR–AUC = 0.41), and that its performance improves with increasing sample size, following a power-law relationship. We utilized an AutoML approach to minimize the risk of overfitting. We further minimized the risk of overly optimistic reporting by using a lock-box set and preregistering our analyses and models. We have made our data, code, and algorithms publicly available on the OSF preregistration site and GitHub. While further external validation is needed, especially regarding the use of pre-trained models for pose estimation, this approach increases confidence in the potential that the pipeline will generalize across datasets, thereby facilitating efforts to share feature datasets and train models across data from multiple sites.

While the GMA has been shown to have a high level of sensitivity and specificity in clinical settings, we did not predict the main important target future outcome – diagnosis of CP – as long-term outcomes were not available at the time of model training. Instead we predicted GMA, a clinician powered risk measure. This is common throughout the automated CP risk prediction literature, with multiple research groups focusing on predicting GMA score, or detecting FMs directly, as opposed to predicting CP diagnosis. This approach is not ideal, as it introduces an additional source of noise from potential human error during assessment, in addition to the noise inherent in the GMA assessment itself. FMs, while highly indicative, are still not a perfect biomarker for CP and multiple items are necessary for CP diagnosis (biomarkers, clinical history, functional motor assessment, and neurological assessment). Over-reliance on FMs risks missing other, perhaps more indicative features or combinations of features that are not readily apparent, and limits risk analyses to the 3–4 month age window. Moreover, the extremely low prevalence of Abnormal FMs makes training a model that captures this movement type infeasible, meaning that some infants at high risk are often not accounted for in models trained only to detect FMs (or their absence). Future efforts should focus directly on predicting CP outcomes, subtypes, and severity.

Our current movement features are likely suboptimal for detecting subtle movement differences. The reason we say this confidently is that others have obtained much better results using approaches optimized to detect FMs. While our features are capturing some differences between groups, they are likely missing some of the subtle movements others have captured with the direct FM featurization. The clinician-selected movement features offer only a coarse description of movement averaged over large windows, whereas we know from the clinical literature that the difference between infants whose movements are typically developing and those that are not is often subtle. For instance, infrequent, small amplitude rolls of the wrists and ankles carry significant clinical meaning, but are infrequent and may be smoothed out when averaged over an entire video. This is especially true when aiming for early prediction before the 3–4-month window. It is also a concern for general-population prescreening, where movement differences may be even more subtle.

Many efforts have been made to identify a precise featurization using machine learning (e.g., [14, 15, 9]). These efforts face inherent challenges due to the small number of FM− infants available for training, which increases the risk of overfitting. The prevalence of infants who develop specific subtypes, or specific levels of severity, is even lower. Pooling data across sites—by sharing a standard set of de-identified features and adopting approaches like AutoML—would enable training on much larger datasets. Models trained on these larger datasets may be able to capture more subtle differences, boosting performance, and enabling even earlier, more precise prediction of CP.

Our current work did not investigate several contextual factors critical for the real-world deployment and performance of predictive models. Future validation must assess the pipeline’s robustness to variations in video quality—such as those from less optimal recording conditions potentially encountered in under-resourced settings—and systematically evaluate the influence of diverse infant skin tones, lighting conditions, and backgrounds on pose estimation accuracy and subsequent predictions. Furthermore, the clinical heterogeneity of CP, which encompasses multiple subtypes and a broad spectrum of severity, is not fully captured by global risk scores like the GMA. A key future direction is therefore to develop models that not only quantify overall risk but also aim to differentiate CP subtypes, leveraging the hypothesis that these distinctions manifest as unique patterns within the movement feature space. Moving towards more nuanced predictions hinges on collecting larger, more diverse datasets with sufficient CP-subtype representation. Achieving this will require large-scale collaborative efforts.

The wide range of ages at which CP is typically diagnosed reflects the fact that less severe movement deficits are often not evident to untrained observers until later in an infant’s development. In contrast, indicators of more severe impairment may be evident to clinicians (and caregivers) much earlier. The infants included in the model all spent time after birth in the Neonatal Intensive Care Unit (NICU), meaning that they were already at an elevated risk of CP. This limitation is prevalent throughout the automated CP detection literature [8, 10, 11, 16], since collecting videos of infants for the purposes of training a ML prediction model is most feasible in a hospital setting. As such the movements that distinguish the two groups in our sample may not be representative of infants from the general infant population. However, other people have shown that movement features can be used to predict GMA scores in at-home videos of infants that are not at high risk [14], so the approach should generalize if trained on the bigger sample that also includes infants from the general population. This should be imminently feasible now that we have released a pose estimation and preprocessing pipeline that is open, easy to share, and does not require fine-tuning.

We have shown that a simple movement-based automated prediction approach performs well in a very large sample. Our model’s performance—ROC-AUC = 0.77 and PR–AUC = 0.41, with powerlaw improvement as sample size increased—should be viewed as particularly encouraging given that it was achieved under rigorous, preregistered conditions, indicating a genuine predictive signal derived from relatively simple features. For any such prescreening tool, the critical challenge lies in balancing sensitivity and specificity: a high false positive rate can unnecessarily worry parents and overburden healthcare systems with healthy children, whereas low sensitivity risks missing children who need clinical intervention, especially in low-resource settings. While our current model demonstrates that achieving a level of predictive accuracy on a large dataset is feasible even under these strict methodological constraints, it clearly requires enhanced precision for broad application, and the present work did not aim to select or evaluate a specific classification threshold. We contend that the most promising path to such improvements is through substantially increasing data scale and diversity by pooling data across many clinical sites. This becomes truly achievable when we prioritize models designed to generalize effectively across different sites and embrace the sharing of de-identified features, an approach our pipeline is built to facilitate.

### Potential implications

One of the biggest limitations in infant CP research is the difficulty in sharing videos due to privacy and safety concerns. There are a growing number of research sites with datasets of over 1000 infant videos, ours is only one example of such a dataset. What is needed is dedicated effort to combine these datasets across sites. While video-based pose estimation for infants is common in the automated detection literature, each site typically uses their own custom, fine-tuned algorithm and a post-processing pipeline tailored to its dataset. This approach hinders broader collaboration and generalizability, though as noted previously, this is increasingly not the case, reflecting a broader shift towards pre-trained models that work across datasets [18, 24].

Here, we present a framework for computing and sharing deidentified features, and training models on the aggregated datasets using AutoML, thereby making it as easy as possible for other researchers to collect large video datasets, compute features, combine across datasets, and train better classifiers. All of the methods used are feasible on a phone camera. All of the videos used for these analyses were collected using handheld iPhones/iPads. The pretrained vision-transformer we used was not fine-tuned on any of the infant videos, and was tested on a wide range of different datasets producing stable results across all of them, as assessed by clinicians. As such we expect that it will work equally well at other clinical sites and on at-home videos. Other researchers have since tested ViTPose-H on infant data and found good performance, and they have released fine-tuned weights specific to infants that they show perform even better [24] and can be specified in the pose-estimation pipeline we provide.

We have shown that advances in pose-estimation now make it feasible to extract movement features from infant videos without the need for any specialized camera setup or fine-tuning. We have shown that movement features derived from these pose estimates predict GMA scores in a very large sample, and that our model has a moderate ROC-AUC (0.77) on the Test (lock-box) set. The simplified process of obtaining de-identified features means that training on datasets across multiple sites and various contexts should now be possible. This facilitates joint efforts and holds tremendous potential for the creation of a global prescreening tool, especially if we boost performance using deep-learned features (including FMs that others have shown perform extremely well at 3-4 months), train on more data including from many infants across sites (powerlaw scaling), and predict CP outcomes as opposed to proxy clinical scores like the GMA.

The capacity to openly share derived movement features (unlike raw videos or detailed keypoints) is crucial. While the development of sophisticated data sharing infrastructures, such as federated learning systems, is a valuable long-term goal, this process often requires considerable time and coordination. To accelerate progress in the interim, our approach leverages features that are not only easy to compute from common video recordings but have also shown predictive power. Their suitability for standardization across multiple clinical sites and developmental stages, combined with their potential for open release, offers an immediate and pragmatic pathway to foster collaboration and build richer aggregated datasets for CP research.

## Methods

### Developing a pipeline for robust skeletal tracking

#### Selecting a pose estimation algorithm

To estimate infant pose from monocular handheld video, we implemented a top-down 2D pose estimation pipeline using tools from the open-source library OpenMMLab. MMDetection was used for infant detection [26], and MMPose was used for 2D pose estimation [26].Infant detection was performed using an RTMDet model [46] pre-trained on the Common Objects in Context (COCO) dataset [47]. Two-dimensional (2D) frame-wise pose estimation was then conducted using ViTPose-H [19], a 10-billion-parameter vision transformer selected for its cross-domain performance.

#### Processing keypoint time series data

Pose estimation was conducted on CHOP high-performance computing servers, ensuring compliance with ethics guidelines by restricting access to CHOP staff and authorized researchers only.

In each frame, only the highest-confidence detection was retained, and frames with keypoint confidence scores below 0.8 were excluded. This process still allowed us to retain over 90% of frames. As in Chambers et al. [5], missing frames were linearly interpolated; outliers were removed with a rolling-median filter (1-second window); and data were smoothed with a rolling-mean filter (1-second window). The effect of smoothing is illustrated in Figure 9.

**Figure 9.**
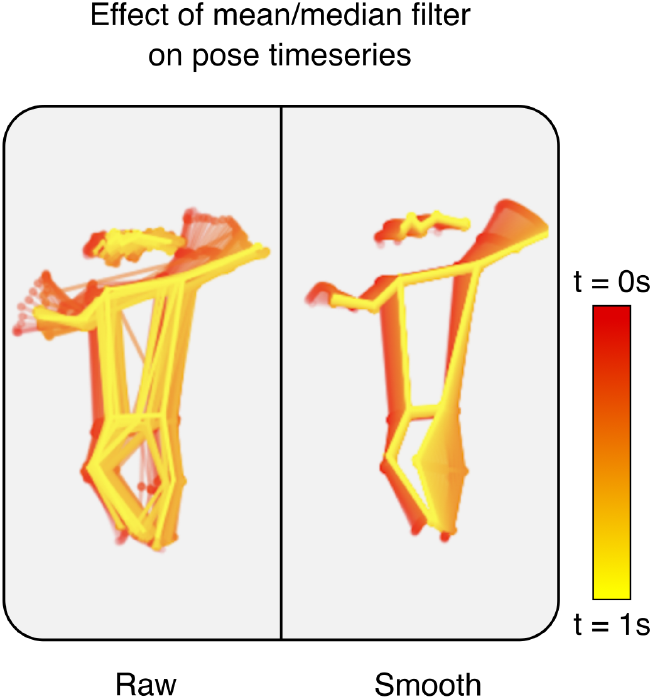
Raw vs smooth pose estimates over a 1s window. Raw time series has inter-frame jitter resulting in noisy time series (left). The mean and median filters (1s) reduce noise (right), but risks losing variability that may be diagnostically significant, particularly in the wrists/ankles.

Future work will include a systematic evaluation of the preprocessing pipeline, particularly the impact of data smoothing. The application and intensity of smoothing techniques can profoundly influence derived kinematic features, potentially obscuring or altering subtle dynamic characteristics critical for distinguishing between movement patterns. While smoothing is a common step to manage noise from pose estimation, the extent to which it affects diagnostically relevant information, such as small-amplitude variability, is not always clear. Therefore, a key future objective will be to systematically quantify these effects. Understanding how different smoothing strategies modify key kinematic outputs will be crucial for optimizing the preprocessing pipeline. This will help preserve clinically meaningful movement details and maximize the analysis’s diagnostic potential.

We chose not to standardize the length of videos, even though it could affect measures of variability (notably standard deviation), for the following reasons. First, an initial check confirmed that the average video duration did not significantly differ between our positive and negative outcome groups, mitigating concerns that video length could act as a systematic confounder at the group level. More importantly, each video was recorded by clinicians with the explicit aim of capturing a sufficient epoch of movement to enable reliable GMA. Thus, the duration of each recording reflects a clinically determined window deemed adequate for observation. The raw standard deviation within such a window directly reflects the movement variability pertinent to this clinical judgment. Normalizing by total frame count might obscure true differences since the proportion of frames containing active, analyzable movement could vary from one clinically sufficient recording to another.

### Dataset split and preregistration

Infant IDs were divided into Train, Validation, and preregistered Test (lock-box) sets using a stratified split to preserve a 10–12% representation of FM–, as well as the ratio of male and female infants and racial and ethnic composition, as described in Table 1. The Test (lock-box) split was defined prior to selection of a pose estimation algorithm and preregistered before any model development or feature computation, ensuring that all downstream analyses were performed without access to the lock-box data. The Train and Val sets were then used within the AutoML framework for model training and internal evaluation, respectively, ensuring that parameter, hyperparameter, and pipeline optimization occurred only on development data. This preregistered design provides a rigorous and unbiased estimate of model generalization for single-site data and represents the strongest available alternative to external multi-site validation.

### Kinematic feature computation

After preregistration and preprocessing, a set of 38 kinematic features were computed from the smoothed keypoint time series using open-source Python code [48] which was adapted from previous work [5]. All features were computed after smoothing the dataset, with frame-wise calculations of joint angles, velocities, and accelerations, which were then aggregated either over the entire video or within overlapping 2-second sliding windows (including rest periods). These features captured displacement, velocity, acceleration, and entropy of the extremities (wrists and ankles) and joint angles (elbows and knees) (Table 2).

No specific features related to GMA FMs were included to minimize the risk of overfitting to the clinical dataset, but may be integrated into future releases.

### Model training

#### Feature selection and preregistration

A binary classifier was trained to predict FM− infants, which indicates a higher risk of developing CP [2, 3, 4, 49, 50]. To reduce the risk of overfitting, feature selection was conducted in consultation with clinicians prior to any data analysis and was preregistered in 2018 [34, 42].Feature computation was automated using custom Python code available on GitHub [48]. Computed features were also preregistered prior to model training.

We did not perform post-hoc iterative feature selection as doing so may lead to overfitting.

#### AutoML framework for model training

Model selection and hyperparameter optimization were carried out using the Auto-sklearn 2.0 package [44, 43], with the “vanilla auto-sklearn” setting (see section: Model training with Auto-sklearn 2.0). Balanced accuracy was chosen as the optimization metric due to the class imbalance (approximately 10:1) [51]. A meta-feature-free portfolio was used for efficient meta-learning, with hyperparameter optimization and model selection performed via successive halving and internal cross-validation within the training data. Six-fold cross-validation was then employed to evaluate model generalizability across different Train/Val splits of the analysis set, and the resulting model was preregistered on June 20, 2024, before being evaluated on the Test (lock-box) set. [34].

## Availability of source code and requirements

- **Project name:** Cerebral palsy risk prediction from video
- **Project homepage:** https://github.com/KordingLab/predict-scores-from-video
- **Operating system(s):** Linux operating system
- **Programming language:** Python
- **Other requirements:** N/A
- **License:** MIT

The source code for skeletal tracking, feature computation, and classifier training has been made available on GitHub and archived on Zenodo [52]. The original feature computation code, from which this work is derived, can be found on GitHub [48].

## Data availability

The dataset supporting the results of this article is available on the OSF repository (https://osf.io/gztmd/) and contains participant IDs, data splits, movement features, and clinical scores [34].

In accordance with ethics guidelines and institutional review board (IRB) approvals from the University of Pennsylvania (Penn) and the Children’s Hospital of Philadelphia (CHOP), study data containing identifiable information are subject to strict handling protocols. Raw videos can only be processed on-site at CHOP and are not shareable externally. Similarly, raw pose-estimates are classified as Protected Health Information (PHI) by the Penn and CHOP IRBs and are therefore not publicly available.

Researchers interested in accessing the raw pose estimate time-series data can request to be added to the IRB protocol by contacting the corresponding author. This process requires completion of mandatory training in HIPAA regulations and relevant clinical research practices. Additional screening by the Penn or CHOP may also be required.

The computed movement features from this study, along with a dataset containing features computed over a 2-second sliding window to preserve temporal information, are available on the project’s OSF preregistration site: osf.io/gztmd/files/osfstorage

For methodological transparency, the pose estimate data from the YouTube-8M subset (94 infants), which was used in selecting our pose estimation algorithm are also available on Figshare [34].

## Data Availability

All data that is consented for sharing (movement features and trained model) are available online at the OSF pre-registration site (https://osf.io/sd6fa)

https://doi.org/10.5281/zenodo.14042732

## List of abbreviations

ROC-AUC: Receiver Operating Characteristic Area Under the Curve
PR-AUC: Precision Recall Area Under the Curve
CHOP: Children’s Hospital of Philadelphia
COCO: Common Objects in Context
CP: Cerebral Palsy
ECMO: Extracorporeal Membrane Oxygenation
FM: Fidgety Movement
FM+: Fidgety Movements Present (GMA Score 1)
FM−: Absent Fidgety Movements (GMA Score 2)
FPR: False Positive Rate
GM: General Movement
GMA: General Movements Assessment
IQR: Inter-Quartile Range
ML: Machine Learning
MRI: Magnetic Resonance Imaging
NICU: Neonatal Intensive Care Unit
TPR: True Positive Rate

## Declarations

### Ethical approval

Ethical approval for this study was provided by the University of Pennsylvania (Penn) Institutional Review Board (IRB Protocol Number: 833180), acting as the single IRB or record and a subsequent reliance agreement between Penn and the Children’s Hospital of Philadelphia (CHOP) Institutional Review Board (IRB Protocol Number: 19-016641).

### Consent for publication

The infant image used in Figure 3 to illustrate algorithm performance is a video frame taken from Adobe Stock Video (ID: #702309262), the use of which is permitted under the Adobe Stock Extended License.

### Author’s contributions

Konrad P. Kording, Michelle J. Johnson, Laura Prosser, and Melanie Segado were responsible for conceptualization of the study aims. Data curation was performed by Andrea F. Duncan, Laura Prosser, and Melanie Segado. Melanie Segado conducted the formal analysis and developed the software. Funding acquisition was led by Konrad P. Kording, Michelle J. Johnson, and Laura Prosser. Data collection and clinical evaluation were carried out by Andrea F. Duncan and Laura Prosser. Methodology was established by Konrad P. Kording and Melanie Segado, with input from Laura Prosser and Michelle J. Johnson. The original draft was written by Melanie Segado and Konrad P. Kording, and all authors contributed to the review and editing of the manuscript.

## Acknowledgements

The authors would like to thank Felipe Parodi for help implementing the pose estimation pipeline, O. Francis Sowande for iterative testing on out-of-sample data, and Yavar Korkian for consistency checking. They would also like to thank Julie Skorup, PT, DPT, PCS and Audrey J Wood, MS, PT, PCS for validation of the skeletal tracking outputs.

